# CARotid plaqUe StabilizatiOn and regression with evolocumab: the CARUSO Study

**DOI:** 10.64898/2026.03.03.26347556

**Authors:** Tiziana Claudia Aranzulla, Andrea Gaggiano, Simone Quaglino, Salvatore Oleandri, Rita D’Aniello, Salvatore Piazza, Marco Pavani, Fabrizio Delnevo, Claudia De Natale, Giuseppe Musumeci

## Abstract

**Background:** Evolocumab promotes coronary plaque regression in patients with coronary artery disease, but little is known regarding carotid plaques (CP). This study aimed to evaluate the impact of evolocumab on top of lipid-lowering therapy (ELLT) on carotid morphological stabilization (MS) and plaque regression (PR) compared to lipid-lowering therapy (LLT) alone.

**Methods:** Asymptomatic patients with internal carotid stenosis≥50% and LDL-C≥100 mg/dL were randomized to ELLT or LLT and monitored by serial duplex ultrasound. The primary endpoint was a composite of 6-month-MS (i.e., switch from morphologic types I-II to III-IV) and/or 12-month-PR (i.e., reduction of carotid stenosis by at least 5% compared to baseline). The secondary endpoint was LDL-C change at 12 months. Major adverse vascular events (MAVE, i.e., cardiac death, stroke, myocardial infarction, carotid or coronary or peripheral revascularization) were recorded.

**Results:** A total of 170 patients were randomized. Mean carotid stenosis was 57%. At 6 months, MS occurred in the ELLT group (10.3%) only (p=0.29). At 12 months, PR was numerically more frequent in the ELLT group, without reaching statistical significance (43% versus 35.1%, p=0.42). The primary endpoint was met in 44.3% versus 35.1% (p=0.26). As compared to baseline, 6 and 12-month shifts from low to high-risk types were significantly higher in the LLT group (p=0.03). The 12-month LDL-C percentage reduction was -73.5% with ELLT, and -48.3% with LLT (p=0.0001). At 1 year, MAVE were significantly more frequent with LLT (14.6% versus 2.4%, p=0.005), and the absence of evolocumab was the only predictor (OR 7, p=0.014).

**Conclusions:** In patients with CP≥50% and LDL-C≥100 mg/dL, ELLT compared to LLT was associated with numerically but not statistically higher 6-month MS and/or 12-month PR. In the LLT group, 6- and 12-month changes from low to high-risk types, LDL-C, and MAVE were significantly higher. According to these results, evolocumab should be considered standard treatment for patients with CP≥50%.

The study was registered at www.clinicaltrials.gov (NCT04730973) and Eudract (2020-005663-31).

**SHORT ABSTRACT:** Patients with carotid stenosis≥50% and LDL-C≥100 mg/dL were randomized to evolocumab on top of optimal lipid-lowering therapy (ELLT) or optimal lipid-lowering therapy (LLT) alone to assess the impact of ELLT on carotid plaque morphological stabilization (MS) and plaque regression (PR). At 6 and 12 months, MS and PR occurred in both groups, but were numerically higher in the ELLT group, without reaching statistical significance. In the LLT group, 6- and 12-month changes from low to high-risk types were significantly higher, and the rate of adverse vascular events was sevenfold higher. Evolocumab might become the standard treatment for patients with carotid artery stenosis ≥50%.

*CLINICAL PERSPECTIVE:* What is new?

- The CARUSO is the largest randomized trial evaluating the impact of evolocumab on top of lipid-lowering therapy (ELLT) on carotid morphological stabilization (MS) and plaque regression (PR) monitored by serial duplex ultrasound.
- The primary endpoint was a composite of 6-month-MS (i.e., switch from morphologic types I-II to III-IV) and/or 12-month-PR (i.e., reduction of carotid stenosis by at least 5% compared to baseline) and was numerically higher in the ELLT group compared to lipid-lowering therapy (LLT) alone, without reaching statistical significance.
- The 1-year rate of major adverse vascular events (MAVE) was sevenfold higher in the LLT group. What are the clinical implications?

- Carotid plaque morphology is a dynamic event, and 6 and 12-month shifts from low to high-risk morphological types were significantly higher in the LLT group, thus suggesting that evolocumab added to LLT may prevent morphological deterioration.
- The absence of evolocumab was the only independent predictor of MAVE; according to our results, ELLT might become the standard treatment for patients with carotid plaques ≥50% and LDL-C not at target.
- Future larger studies are warranted to validate our findings, assess long-term adherence to therapy, and identify subgroups with higher probability of achieving MS and PR.

## INTRODUCTION

Lipid-lowering therapy (LLT) is a therapeutic pillar for the management of atherosclerotic vascular disease. Indeed, major adverse vascular events (MAVE) and progression of atherosclerosis are reduced in proportion to the achieved LDL cholesterol (LDL-C) levels, with no LDL-C lower threshold below which the benefit ceases or harm occurs.^1–3^ In particular, optimal LLT (which causes a LDL-C reduction ≥50% from baseline) has been associated with regression of atherosclerotic plaques.^4^ Current guidelines recommend LDL-C values <55 mg/dL in patients at very high risk, as those with documented - i.e., on carotid Duplex ultrasound (DUS) - atherosclerotic cardiovascular disease.^5^ However, LLT may be limited by side effects (i.e., disabling myalgia) which are dose-dependent. Furthermore, the maximum tolerated statin dose might still be insufficient to reach the recommended LDL-C goals. The advent of proprotein convertase subtilisin kexin type 9 inhibitors (PCSK9-inhibitors) overcame these limitations. Therefore, when the maximum tolerated LLT is not sufficient, the addition of PCSK9-inhibitors is indicated.^5, 6^ In patients with coronary artery disease (CAD), therapy with PCSK9-inhibitors has been associated with favorable outcomes,^2, 7, 8^ and coronary plaque regression.^9–11^ Conversely, little is known regarding carotid plaque (CP) modification. Case reports^12^ and very small randomized studies showed carotid morphological stabilization (MS)^13^ and plaque regression (PR),^14^ with evolocumab.

The CARotid plaqUe StabilizatiOn and regression with evolocumab (CARUSO) study aimed at evaluating carotid MS and PR following therapy with evolocumab on top of LLT.

## METHODS

The rationale and design of the CARUSO study have been previously detailed.^15^ Briefly, in this single-center investigator-initiated trial, asymptomatic patients with mono- or bilateral de novo CP≥50% according to the ESCT criteria at DUS and LDL-C values≥100 mg/dL were randomized to either evolocumab on top of LLT (ELLT) or optimized LLT alone, that is, rosuvastatin≥20 mg and ezetimibe (Figure 1). Randomization was performed immediately after DUS, and therapy was started the same day. All the patients on other statins were switched to the study LLT. In the ELLT group, biweekly evolocumab 140 mg was administered s.c. The study allocation was not revealed to the investigators. Although patients randomized to LLT did not receive a subcutaneous placebo, they were not told that the comparator drug was administered subcutaneously.

**Figure 1.**
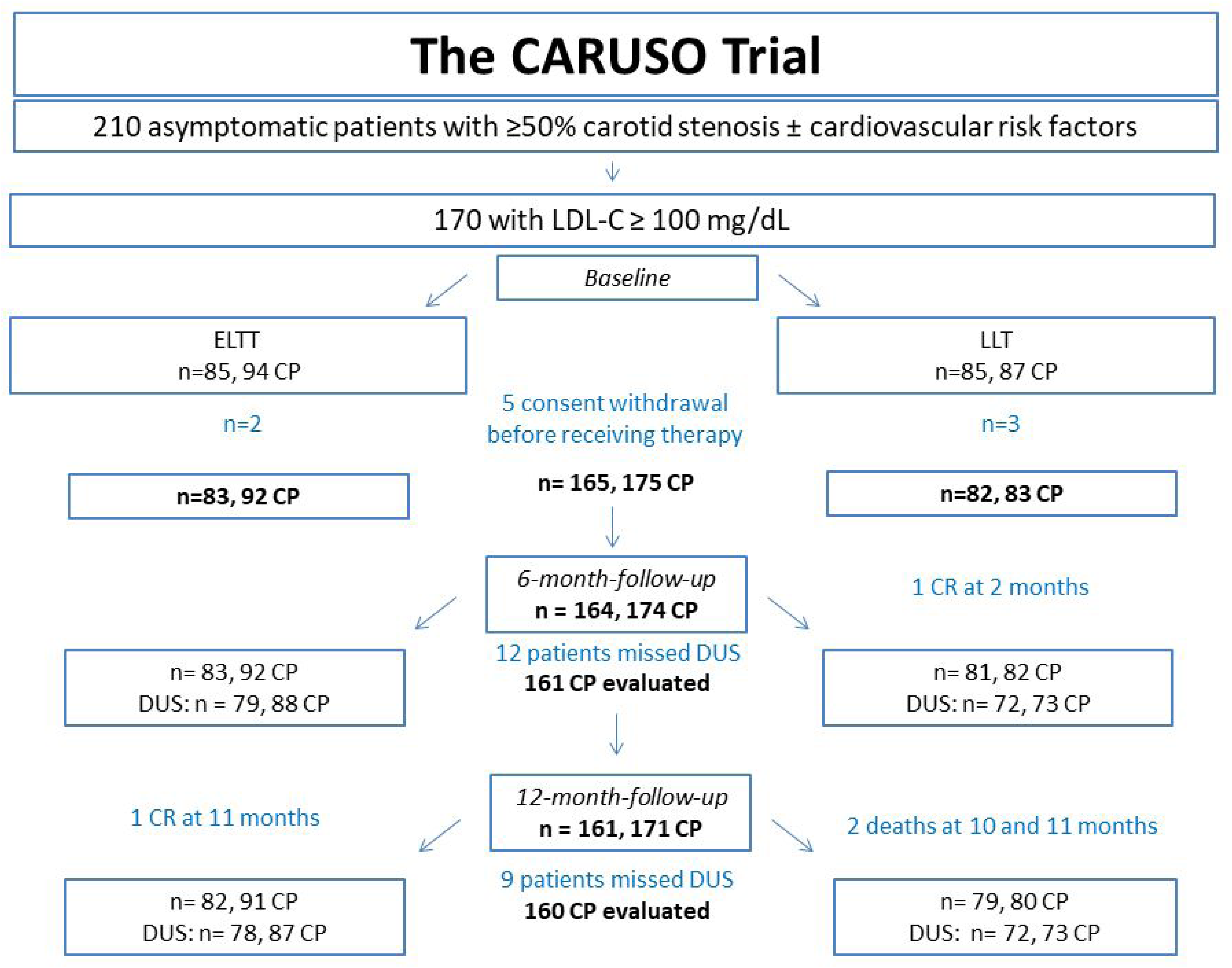
CONSORT diagram of the CARUSO (CARotid plaqUe StabilizatiOn and regression with evolocumab) Study. LLT= optimal lipid-lowering therapy; ELLT= evolocumab on top of LLT. Carotid plaques = CP, DUS= duplex ultrasound, CR= carotid revascularization

Baseline cardiovascular risk factors, LDL-C values, renal function, glycemic status, and overall ongoing therapies, including antiplatelets and statins before randomization were recorded. In addition to compliance with medical therapy, a healthy lifestyle, physical activity, and blood pressure control were recommended. The study was conducted under the ethical principles contained in the Declaration of Helsinki and the local ethics committee approved it. Each patient provided signed informed consent. The number of patients screened, randomized, treated, and analyzed is reported according to the CONSORT guidelines. The study was registered at www.clinicaltrials.gov (NCT04730973) and Eudract (2020-005663-31).

### Exclusion criteria

Patients aged <18 or >80 years, with known intolerance to evolocumab, ongoing or previous treatment with PCSK9-inhibitors, stroke or transient ischemic attack (TIA) within 12 months, complete carotid occlusion, or unwilling to provide informed consent were excluded. Patients with previous carotid revascularization (CR) were included if the contralateral carotid had a de novo≥50% stenosis.

### Examinations

Baseline morphological features of the internal carotid artery were carefully assessed and CP were graded as type I, II, III, IV based on increasing echogenicity, type I: predominantly hypoechoic plaque with thin echogenic rim; type II: echogenic plaque with >50% hypoechoic areas; type III: echogenic plaque with <50% hypoechoic areas; type IV: uniformly echogenic plaque^16^(Figure 2). As detailed in the study protocol,^15^ stenosis severity was assessed with longitudinal and cross-sectional B-mode and color Doppler and ESCT criteria were used; peak systolic velocity (PSV) and end-diastolic velocity (EDV) were routinely evaluated. All DUS were performed at our center by two experienced vascular surgeons who were blinded to the therapeutic regimen. They separately performed each DUS. When discordance on the morphological type or measures was found, consensus was achieved involving a third vascular surgeon. Carotid DUS was repeated at 6 and 12 months.

**Figure 2.**
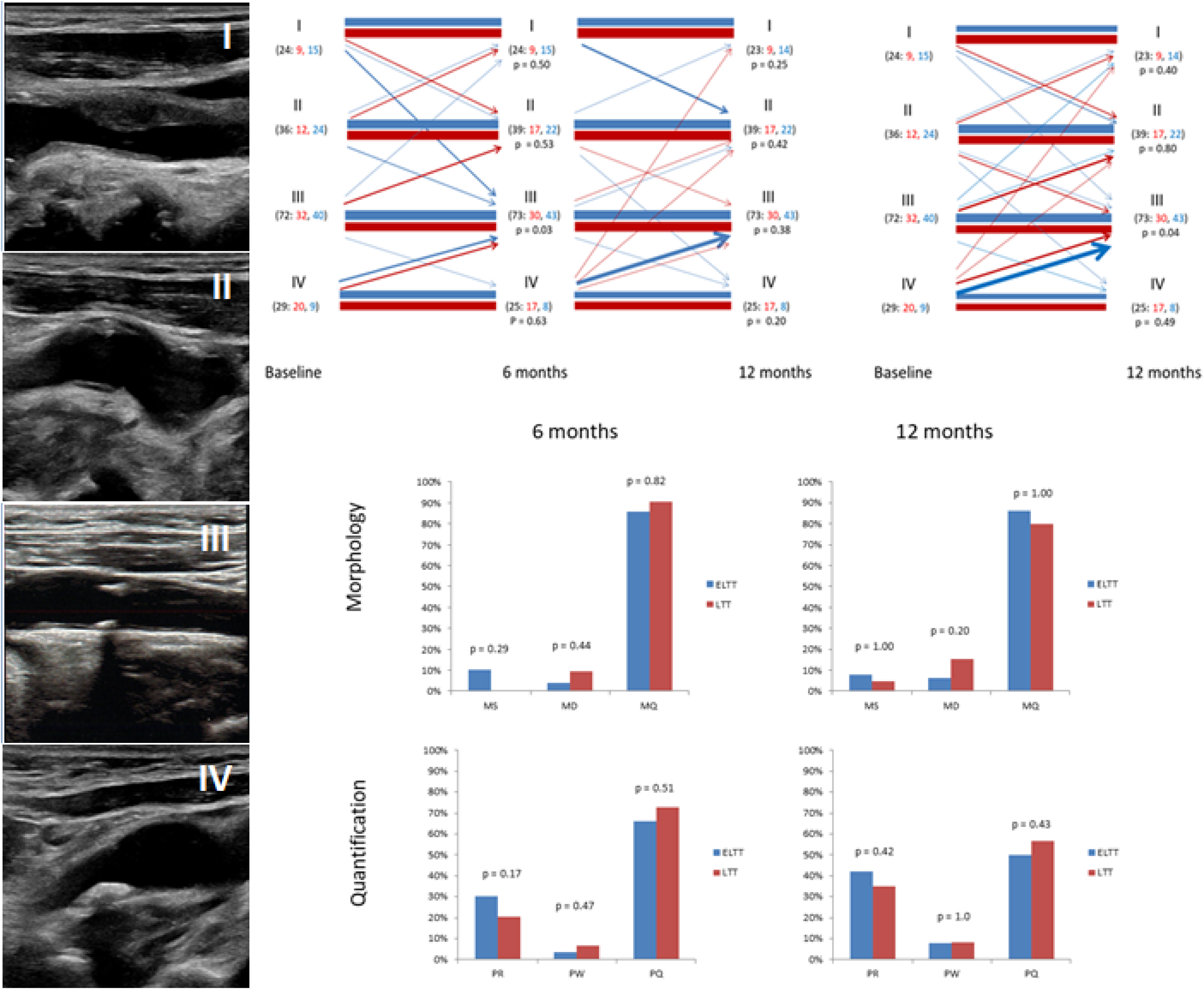
In vertical: Carotid plaque morphological types at Doppler UltraSound. I: predominantly hypoechoic plaque with a thin echogenic rim, II: echogenic plaque with >50% hypoechoic areas, III: echogenic plaque with <50% hypoechoic areas, IV: uniformly echogenic plaque. In horizontal, higher panel: Morphological changes at different timepoints. Line depths are proportional to the percentages of shifts. Number in brackets: in black, the overall number of plaques; in red, the number of LLT plaques; in blue, the number of ELLT plaques. Lower panel: Morphological and Quantitative changes at 6 and 12 months. LLT= optimal lipid-lowering therapy; ELLT= evolocumab on top of LLT. MS = morphological stabilization, MD = morphological deterioration, MQ = morphological quiescence, PR = plaque regression, PW = plaque worsening, PC = plaque quiescence

Blood samples were collected at baseline, 6, and 12 months in the morning under fasting conditions. When possible, LDL-C was requested as a direct measurement.

### Study endpoint

The primary endpoint was the superiority of ELLT versus LLT regarding MS at 6 months and/or PR at 12 months.

Morphological stabilization was defined as the shift from high-risk types (I or II) to low-risk (III or IV) types with the achievement of a regular plaque morphology and prevalent fibrous atheroma (Figure 2). Carotid PR was defined as a decrease in stenosis degree by at least 5%, compared to baseline. The secondary endpoint was the absolute and percentage LDL-C change at 12 months. Also, 12-month-MS, and morphologic deterioration (MD, i.e., the proportion of low-risk plaques that shifted to high-risk types, that is the opposite of MS), or morphologic quiescence (MQ, i.e., the absence of any change in the morphological risk class) at 6 and 12 months were documented. Similarly, plaque worsening (PW, i.e., any increase of stenosis degree) and plaque quiescence (PQ, i.e., the absence of any change in stenosis degree). These concepts and definitions are summarized in Table I.

**Table I.** Carotid plaque modifications and study end-points.

| Event | Definition |
| --- | --- |
| *Carotid Plaque Morphology* |  |
| Morphological Stabilization (MS) | Switch from high-risk (I-II) to low-risk (III-IV) types, that is the achievement of a regular plaque morphology with prevalence of fibrous atheroma |
| Morphologic deterioration (MD) | Switch from low-risk (III-IV) to high-risk (I-II) types, that is the opposite of morphologic stabilization |
| Morphologic quiescence (MQ) | Absence of any shift from the previous morphological risk class |
| *Carotid Plaque Entity* |  |
| Plaque Regression (PR) | Reduction of stenosis degree by at least 5% compared to baseline |
| Plaque worsening (PW) | Any increase of stenosis degree |
| Plaque Quiescence (PQ) | Absence of any change in stenosis degree |
| Primary endpoint | MS at 6 months and/or PR at 12 months |
| Secondary endpoint | Absolute and percentage LDL-C variation at 12 months |
| Major Vascular Adverse Events (MAVE) | Cardiac death, stroke, myocardial infarction, carotid or coronary or peripheral revascularization |
| *Therapy* |  |
| Lipid-lowering therapy (LLT) | Rosuvastatin 20 mg + Ezetimibe 10 mg daily |
| Evolocumab added to lipid-lowering therapy (ELLT) | Evolocumab 140 mg every two weeks and Rosuvastatin 20 mg + Ezetimibe 10 mg daily |

### Adverse events and study conclusion

All-cause mortality and MAVE (including cardiac death, stroke, myocardial infarction, carotid or coronary or peripheral revascularization) were collected by outpatient visits, telephone follow-up, and/or contact with treating physicians. Similarly, any adverse event, including new-onset diabetes, cognitive impairment or neurological disease, and cancer, was reported.

In case of serious adverse events (i.e., allergic reactions or gastroenteric /hepatic intolerance to evolocumab) or consent withdrawal, patients dropped out of the study. Patients concluded the study if CR (which was considered a missed primary endpoint) or death occurred.

### Statistical analysis

Continuous variables were compared with T-test if normally distributed (mean±standard deviation) and with Mann-Whitney U if not normally distributed (median; interquartile range). For categorical variables (frequencies), the chi-square test was used. Six- and 12-month data were compared with baseline data in each group with paired T-test or paired Mann-Whitney U and chi-square. Between-groups and within-groups differences were calculated with two-way mixed-design repeated measures ANOVA for continuous variables and logistic regression for categorical variables. Multivariate regression analyses were performed to identify independent predictors of the primary endpoint and of hierarchically assessed MAVE. Specifically, variables that were significantly associated with the event on univariate analysis were entered into multivariate Cox proportional-hazards models, and analyses were performed with the enter method. The Hosmer-Lemeshow goodness-of-fit test was used to validate the model. We reported odds ratios (OR) and associated 95% confidence intervals (CI). The assessment of time-to-event clinical outcomes was done using Kaplan-Meier survival curves, which were compared with the log-rank test.

For all statistical analyses, a 2-sided p-value <0.05 was considered significant; SPSS version 31 (IBM) for Windows was used.

## RESULTS

A total of 170 patients with 181 CP ≥50% were randomized to ELLT or LLT. The first patient was enrolled in October 2023, the last in April 2024. Twelve-month follow-up was completed in May 2025. Five patients (one with 2 CP) withdrew consent before receiving either LLT or ELLT. The study analysis included 165 patients with 175 CP (Figure 1).

### Baseline characteristics

Baseline clinical characteristics are shown in Table II. Mean age was 72 years, 61% were males, and 40% had diabetes. A previous MI or stroke occurred in 16% and 4%, respectively. Antiplatelet therapy was taken at baseline by 81%; it was prescribed in all after enrollment. The two groups were well balanced. In 37% DUS was indicated by a cardiologist, in 33% by a vascular surgeon, and in 30% by a diabetologist. DUS was available for 151 patients with 161 plaques at 6 months and for 150 patients with 160 plaques at 12 months (Figure 1).

**Table II.**
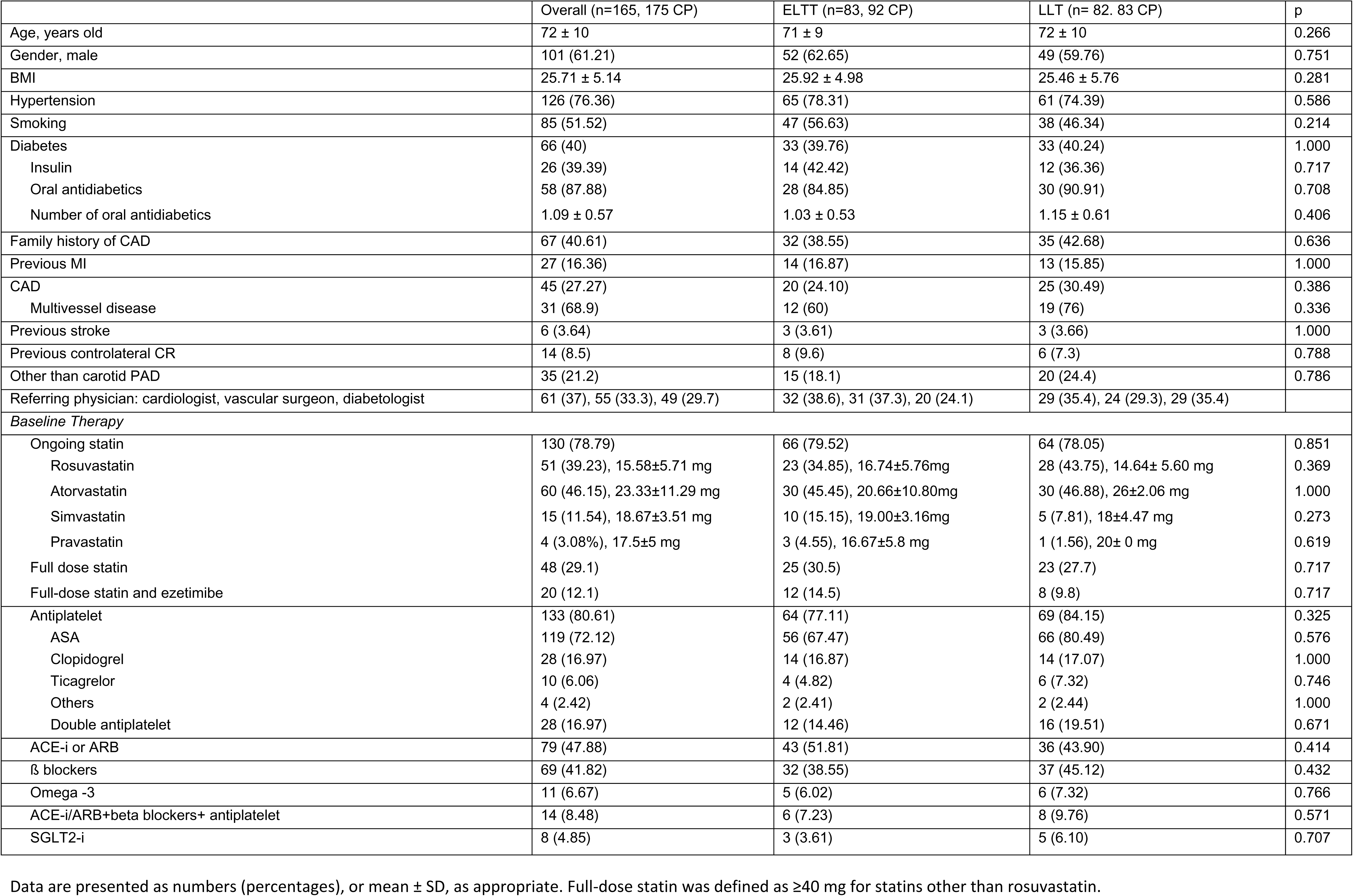
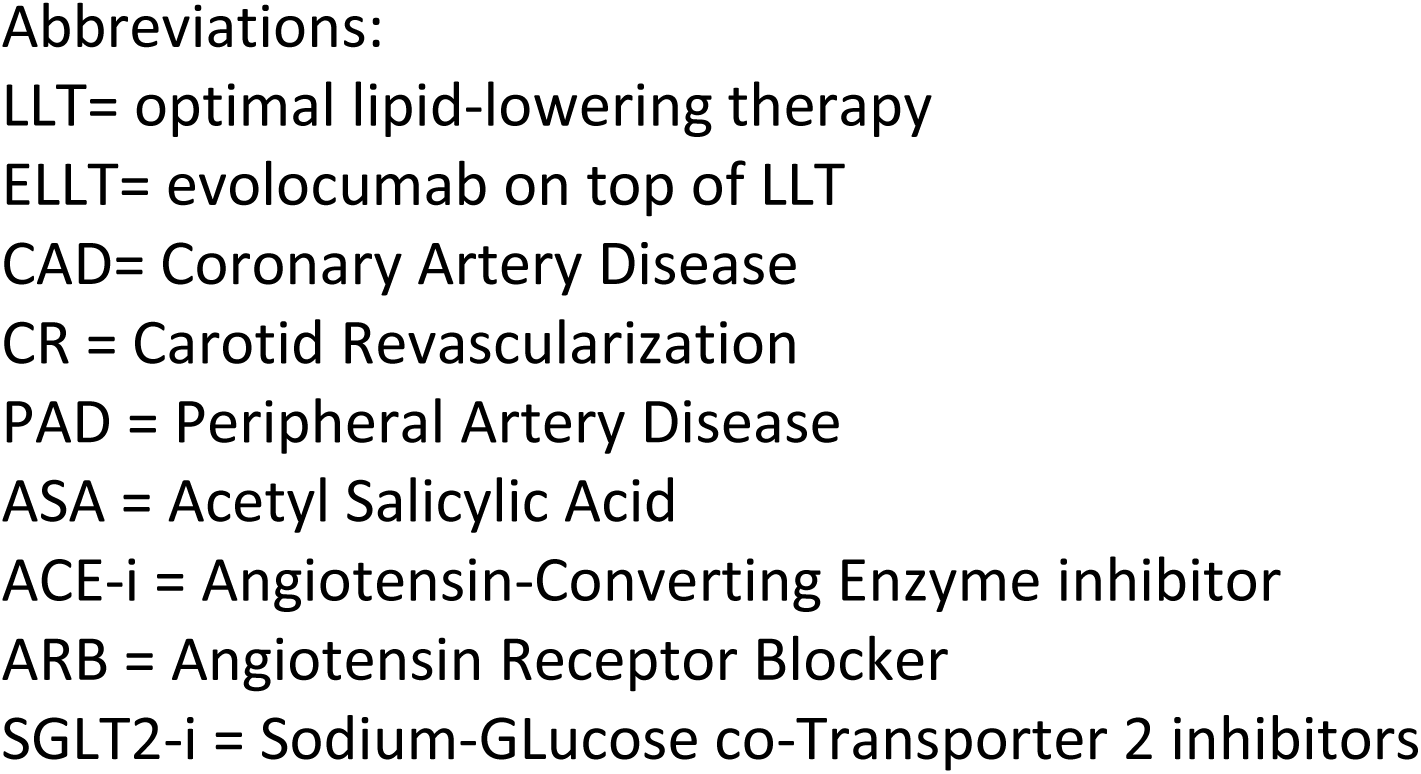
Baseline characteristics of the study populations.

|  | Overall (n=165, 175 CP) | ELTT (n=83, 92 CP) | LLT (n= 82. 83 CP) | p |
| --- | --- | --- | --- | --- |
| Age, years old | 72 ± 10 | 71 ± 9 | 72 ± 10 | 0.266 |
| Gender, male | 101 (61.21) | 52 (62.65) | 49 (59.76) | 0.751 |
| BMI | 25.71 ± 5.14 | 25.92 ± 4.98 | 25.46 ± 5.76 | 0.281 |
| Hypertension | 126 (76.36) | 65 (78.31) | 61 (74.39) | 0.586 |
| Smoking | 85 (51.52) | 47 (56.63) | 38 (46.34) | 0.214 |
| Diabetes | 66 (40) | 33 (39.76) | 33 (40.24) | 1.000 |
| Insulin | 26 (39.39) | 14 (42.42) | 12 (36.36) | 0.717 |
| Oral antidiabetics | 58 (87.88) | 28 (84.85) | 30 (90.91) | 0.708 |
| Number of oral antidiabetics | 1.09 ± 0.57 | 1.03 ± 0.53 | 1.15 ± 0.61 | 0.406 |
| Family history of CAD | 67 (40.61) | 32 (38.55) | 35 (42.68) | 0.636 |
| Previous MI | 27 (16.36) | 14 (16.87) | 13 (15.85) | 1.000 |
| CAD | 45 (27.27) | 20 (24.10) | 25 (30.49) | 0.386 |
| Multivessel disease | 31 (68.9) | 12 (60) | 19 (76) | 0.336 |
| Previous stroke | 6 (3.64) | 3 (3.61) | 3 (3.66) | 1.000 |
| Previous controlateral CR | 14 (8.5) | 8 (9.6) | 6 (7.3) | 0.788 |
| Other than carotid PAD | 35 (21.2) | 15 (18.1) | 20 (24.4) | 0.786 |
| Referring physician: cardiologist, vascular surgeon, diabetologist | 61 (37), 55 (33.3), 49 (29.7) | 32 (38.6), 31 (37.3), 20 (24.1) | 29 (35.4), 24 (29.3), 29 (35.4) |  |
| *Baseline Therapy* |  |  |  |  |
| Ongoing statin | 130 (78.79) | 66 (79.52) | 64 (78.05) | 0.851 |
| Rosuvastatin | 51 (39.23), 15.58±5.71 mg | 23 (34.85), 16.74±5.76mg | 28 (43.75), 14.64± 5.60 mg | 0.369 |
| Atorvastatin | 60 (46.15), 23.33±11.29 mg | 30 (45.45), 20.66±10.80mg | 30 (46.88), 26±2.06 mg | 1.000 |
| Simvastatin | 15 (11.54), 18.67±3.51 mg | 10 (15.15), 19.00±3.16mg | 5 (7.81), 18±4.47 mg | 0.273 |
| Pravastatin | 4 (3.08%), 17.5±5 mg | 3 (4.55), 16.67±5.8 mg | 1 (1.56), 20± 0 mg | 0.619 |
| Full dose statin | 48 (29.1) | 25 (30.5) | 23 (27.7) | 0.717 |
| Full-dose statin and ezetimibe | 20 (12.1) | 12 (14.5) | 8 (9.8) | 0.717 |
| Antiplatelet | 133 (80.61) | 64 (77.11) | 69 (84.15) | 0.325 |
| ASA | 119 (72.12) | 56 (67.47) | 66 (80.49) | 0.576 |
| Clopidogrel | 28 (16.97) | 14 (16.87) | 14 (17.07) | 1.000 |
| Ticagrelor | 10 (6.06) | 4 (4.82) | 6 (7.32) | 0.746 |
| Others | 4 (2.42) | 2 (2.41) | 2 (2.44) | 1.000 |
| Double antiplatelet | 28 (16.97) | 12 (14.46) | 16 (19.51) | 0.671 |
| ACE-i or ARB | 79 (47.88) | 43 (51.81) | 36 (43.90) | 0.414 |
| β blockers | 69 (41.82) | 32 (38.55) | 37 (45.12) | 0.432 |
| Omega -3 | 11 (6.67) | 5 (6.02) | 6 (7.32) | 0.766 |
| ACE-i/ARB+beta blockers+ antiplatelet | 14 (8.48) | 6 (7.23) | 8 (9.76) | 0.571 |
| SGLT2-i | 8 (4.85) | 3 (3.61) | 5 (6.10) | 0.707 |

Data are presented as numbers (percentages), or mean ± SD, as appropriate. Full-dose statin was defined as ≥40 mg for statins other than rosuvastatin.
Abbreviations: LLT= optimal lipid-lowering therapy ELLT= evolocumab on top of LLT CAD= Coronary Artery Disease CR = Carotid Revascularization PAD = Peripheral Artery Disease ASA = Acetyl Salicylic Acid ACE-i = Angiotensin-Converting Enzyme inhibitor ARB = Angiotensin Receptor Blocker SGLT2-i = Sodium-GLucose co-Transporter 2 inhibitors

### Plaques

At baseline, plaque characteristics were equally distributed between the 2 groups (Table III), except for a higher prevalence of high-risk plaques in the EOLLT group (44.6 versus 28.9, p=0.04). This basal difference was mostly driven by the lowest-risk type IV and persisted after consideration of only those CP with a 6-month follow-up DUS.

**Table III.** Carotid plaque characteristics during the study.

| Outcome | Timepoint | N | Overall |  | N | ELLT |  | N | LLT |  |  | Between-Groups | Within-Group | Interaction |
| --- | --- | --- | --- | --- | --- | --- | --- | --- | --- | --- | --- | --- | --- | --- |
|  |  |  | Values | Change |  | Values | Change |  | Values | Change | Mean (95% CI) | p | p | p |
| High-risk type (I + II) | Baseline | 175 | 65 (37) |  | 92 | 41 (45) |  | 83 | 24 (29) |  |  | 0.04 |  |  |
|  | Baseline CP with 6 month-DUS | 161 | 60 (37) |  | 88 | 39 (44) |  | 73 | 21 (29) |  |  | 0.05 |  |  |
|  | 6 months vs baseline | 161 | 63 (39) | +3 | 88 | 37 (42) | -2 | 73 | 26 (36) | +5 |  | 0.03 | 0.05 | 0.043 |
|  | 6 month-CP with 12 month-DUS | 160 | 62 (39) |  | 87 | 36 (41) |  | 73 | 26 (36) |  |  |  |  |  |
|  | 12 months vs 6 months | 160 | 66 (41) | +4 | 87 | 38 (44) | +2 | 73 | 28 (38) | +2 |  | 0.408 | 0.319 | 0.761 |
|  | 12 months vs baseline | 160 | 66 (41) | +6 | 87 | 38 (44) | -1 | 73 | 28 (38) | +7 |  | 0.03 | 0.027 | 0.050 |
| Type I | Baseline | 175 | 27 (15) |  | 92 | 16 (17) |  | 83 | 11 (13) |  |  | 0.532 |  |  |
|  | Baseline CP with 6 month-DUS | 161 | 24 (15) |  | 88 | 15 (17) |  | 73 | 9 (12) |  |  | 0.506 |  |  |
|  | 6 months vs baseline | 161 | 24 (15) | 0 | 88 | 15 (17) | 0 | 73 | 9 (12) | 0 |  | 0.450 | 0.706 | 0.879 |
|  | 6 month-CP with 12 month-DUS | 160 | 23 (14) |  | 87 | 14 (16) |  | 73 | 9 (12) |  |  |  |  |  |
|  | 12 months vs 6 months | 160 | 22 (14) | -1 | 87 | 12 (14) | -2 | 73 | 10 (14) | +1 |  | 0.401 | 0.319 | 0.105 |
|  | 12 months vs baseline | 160 | 22 (14) | -2 | 87 | 12 (14) | -3 | 73 | 10 (14) | +1 |  | 0.450 | 0.874 | 0.362 |
| Type II | Baseline | 175 | 38 (22) |  | 92 | 25 (27) |  | 83 | 13 (16) |  |  | 0.070 |  |  |
|  | Baseline CP with 6 month-DUS | 161 | 36 (22) |  | 88 | 24 (27) |  | 73 | 12 (16) |  |  | 0.129 |  |  |
|  | 6 months vs baseline | 161 | 39 (24) | +3 | 88 | 22 (25) | -2 | 73 | 17 (23) | +5 |  | 0.063 | 0.039 | 0.027 |
|  | 6 month-CP with 12 month-DUS | 160 | 39 (24) |  | 87 | 22 (25) |  | 73 | 17 (23) |  |  |  |  |  |
|  | 12 months vs 6 months | 160 | 44 (28) | +5 | 87 | 26 (30) | +4 | 73 | 18 (25) | +1 |  | 0.802 | 0.567 | 0.383 |
|  | 12 months vs baseline | 160 | 44 (28) | +8 | 87 | 26 (30) | +2 | 73 | 18 (25) | +6 |  | 0.063 | 0.037 | 0.250 |
| Type III | Baseline | 175 | 80 (46) |  | 92 | 42 (46) |  | 83 | 38 (46) |  |  | 1.000 |  |  |
|  | Baseline CP with 6 month-DUS | 161 | 73 (45) |  | 88 | 40 (45) |  | 73 | 33 (45) |  |  | 1.000 |  |  |
|  | 6 months vs baseline | 161 | 73 (45) | 0 | 88 | 43 (49) | +3 | 73 | 30 (41) | -3 |  | 0.986 | 0.266 | 0.151 |
|  | 6 month-CP with 12 month-DUS | 160 | 73 (46) |  | 87 | 43 (49) |  | 73 | 30 (41) |  |  |  |  |  |
|  | 12 months vs 6 months | 160 | 72 (45) | -1 | 87 | 41 (47) | -2 | 73 | 31 (42) | +1 |  | 0.327 | 0.567 | 0.446 |
|  | 12 months vs baseline | 160 | 72 (45) | -1 | 87 | 41 (47) | +1 | 73 | 31 (42) | - 2 |  | 0.986 | 0.488 | 0.439 |
| Type IV | Baseline | 175 | 30 (17) |  | 92 | 9 (10) |  | 83 | 21 (25) |  |  | 0.009 |  |  |
|  | Baseline CP with 6 month-DUS | 161 | 29 (18) |  | 88 | 9 (10) |  | 73 | 20 (27) |  |  | 0.007 |  |  |
|  | 6 months vs baseline | 161 | 25 (16) | -4 | 88 | 8 (9) | -1 | 73 | 17 (23) | -3 |  | 0.007 | 0.434 | 0.682 |
|  | 6 month-CP with 12 month-DUS | 160 | 25 (16) |  | 87 | 8 (9) |  | 73 | 17 (23) |  |  |  |  |  |
|  | 12 months vs 6 months | 160 | 22 (14) | -3 | 87 | 8 (9) | 0 | 73 | 14 (19) | -3 |  | 0.016 | 0.134 | 0.909 |
|  | 12 months vs baseline | 160 | 22 (14) | -7 | 87 | 8 (9) | -1 | 73 | 14 (19) | -6 |  | 0.007 | 0.441 | 0.903 |
| Carotid stenosis, % | Baseline | 175 | 57.04 ± 5.24 |  | 92 | 57.44 ± 5.45 |  | 83 | 56.59 ± 5 |  |  | 0.283 |  |  |
|  | 6 months vs baseline | 161 | 55.74 ± 6.26 | -1.50 ± 3.93 | 88 | 55.59 ± 6.47 | -1.91 ± 4.44 | 73 | 55.93 ± 6.03 | -1.01 ±3.6 | 0.895 (-0.33 2.12) | 0.629 | 0.0001 | 0.150 |
|  | 12 months vs 6 months | 160 | 55.23 ± 7.00 | -0.45 ±3.82 | 87 | 55.32 ± 7.56 | -0.16 ± 4.58 | 73 | 55.14 ± 6.33 | -0.79±2.66 | -0.633 (-1.83 0.56) | 0.981 | 0.117 | 0.294 |
|  | 12 months vs baseline | 160 | 55.23 ± 7.00 | -1.96 ±5.18 | 87 | 55.32 ± 7.56 | -2.07 ± 6.20 | 73 | 55.14 ± 6.33 | -1.80±3.66 | 0.28 (-1.34 1.91) | 0.412 | 0.000 | 0.731 |
| Stenosis reduction > 5% | 6 months vs baseline | 161 | 43 (27) |  | 88 | 28 (32) |  | 73 | 15 (21) |  |  | 0.105 | 0.854 | 0.172 |
|  | 12 months vs 6 months | 160 | 33 (21) | -10 | 87 | 17 (20) | -11 | 73 | 16 (22) | +1 |  | 0.688 | 0.006 | 0.157 |
|  | 12 months vs baseline | 160 | 63 (39) |  | 87 | 37 (43) |  | 73 | 26 (36) |  |  | 0.105 | 0.003 | 0.527 |
| PSV | Baseline | 175 | 125.46 ± 55 |  | 92 | 127.85 ± 60 |  | 83 | 122.81 ± 43.41 |  |  | 0.723 |  |  |
|  | 6 months vs baseline | 159 | 121.40 ± 46.50 | -3.42 ±24.48 | 86 | 123.77 ± 49.42 | -2.41 ± 25.64 | 73 | 118.60 ± 45.04 | -4.61 ± 23.86 | -2.21 (-9.92, 5.50) | 0.372 | 0.073 | 0.572 |
|  | 12 months vs 6 months | 159 | 120.90 ± 44.63 | -0.50 ±32.81 | 86 | 126.40 ± 44.41 | 0.66 ± 39.44 | 73 | 116.72 ± 45.17 | -1.87 ± 22.86 | -2.53 (-12.87, 4.63) | 0.356 | 0.816 | 0.628 |
|  | 12 months vs baseline | 159 | 120.90 ± 48.72 | -3.92 ±36.65 | 86 | 124.44 ± 54.54 | -1.74 ± 45.09 | 73 | 116.72 ± 45.17 | -6.49 ± 23.10 | -4.74 (-16.28, 6.78) | 0.281 | 0.160 | 0.417 |
| PSV reduction > 5% | 6 months vs baseline | 159 | 72 (45) |  | 86 | 37 (43) |  | 73 | 35 |  |  | 0.538 | 0.084 | 0.764 |
|  | 12 months vs 6 months | 159 | 51 (32) | -21 | 86 | 27 (32) | -10 | 73 | 24 | -11 |  | 0.844 | 0.009 | 0.579 |
|  | 12 months vs baseline | 159 | 73 (46) |  | 86 | 37 (43) |  | 73 | 36 |  |  | 0.538 | 0.820 | 0.854 |

Data are presented as numbers (percentages), or mean ± SD, as appropriate
Numeric changes are the net algebraic changes and were calculated considering only available data
Example: High-risk type carotid plaques (CP)
- Overall CP at baseline were 175. Overall plaques with baseline and 6-month Doppler UltraSound (DUS) were 161. Of these, 60 were high-risk at baseline; they were 63 at 6 months. The net change is +3. - Overall CP with DUS at both 6 and 12 months were 160. Of these, 62 were high-risk at 6 months; they were 66 at 12 months. The net change is +4. - Overall CP with baseline, 6 and 12 month-DUS were 160. At baseline there were 60 high-risk CP, at 12 months 66. The net change is +6.
N are the CP with data available for each time point
Between-groups: effect of time periods (6months versus baseline, 12 months versus 6 months, 12 months versus baseline) across the two groups
Within-group: effect of therapy across the time periods in each group; i.e., effect of EOLLT across the time periods in the EOLLT group
Interaction: indicates whether the effect of therapy depends on the time periods
LLT= optimal lipid-lowering therapy
ELLT= evolocumab on top of LLT

#### Morphological changes

Morphological variations of CP over time occurred in both directions, including shifts from low to high-risk types and vice versa. These changes are reported in Table III.

Overall, the net number of high-risk CP increased over time, with a net algebraic increase of +3 high risk plaques at 6 months, +4 between 6 and 12 months, and a net balance of +6 at 12 months compared to baseline

Within the ELLT group, the net number of high-risk plaques decreased at 6 months (−2), then increased from 6 to 12 months (+2).

Within the LLT group, morphological changes over time determined a net increase of the number of high-risk plaques at each timepoint (+5 from baseline to 6 months, p=0.05; +2 between 6 and 12 months, p = 0.32; +7 from baseline to 12 months, p=0.027). Most of these shifts occurred to type II. Between the two treatment groups, the net shifts from low to high-risk CP was significantly higher with LLT compared to ELLT at both 6 (p =0.03) and 12 months (p =0.03) compared to baseline.

During the period between 6 and 12 months, no significant variation in morphologic shifts from low-to high-risk types occurred within each group (p=0.32). Similarly, no significant difference between the ELLT and LLT groups was detected regarding these variations (p=0.41), suggesting that morphological changes occurred mostly at 6 months, and the 12–month significant difference compared to baseline was likely or mostly driven by the 6 month-changes.

Changes between morphological types from baseline to 6 months, from 6 to 12 months, and from baseline to 12 months are shown in Figure 2. Type III was the most dynamic morphological type. From baseline to 6 months, out of 32 type III CP, 5 (15.6%) worsened to type II in the LLT group, while in the ELLT group, out of 40 type III CP, 7.5% shifted (p =0.03): 2 (5%) worsened to type I and 1 (2.5%) improved to type IV. From baseline to 12 months, 6 (18.8%) type III CP worsened to type II in the LLT group, while in the ELLT group, 12.5% type III CP shifted (p =0.04): 2 (5%) shifted to type I, 1 (2.5%) to type II, and 2 (5%) to type IV. Changes of type III were mostly driven by the 6-month-changes.

Morphological variations are depicted in Figure 2.

Among 60 high-risk CP at baseline with available 6-month DUS (39 ELLT and 21 LLT), MS occurred in 4 (10.26%), all within the ELLT group (p=0.29). In the subset of high-risk CP, 12-month MS was found in 4 plaques: 2 maintained 6-month MS, and other 2 acquired MS thereafter, while the 2 CP that had acquired 6-month MS deteriorated at 12 months.

Among 101 low-risk CP at baseline with available 6-month DUS (49 ELLT and 52 LLT), MD occurred in 7 plaques (2 ELLT and 5 LLT, p =0.44). At 12 months, MD was found in 11 plaques (3 ELLT and 8 LLT). Of those, 7 (2 ELLT, 5 LLT) had already deteriorated at 6 months, 4 (1 ELLT, 3 LLT) deteriorated thereafter. Overall, MQ occurred in 87% and 80% at 6 and 12 months, respectively, with no statistical differences between the two groups. When joining MS and MQ, they involved 92% ELLT and 86.3% LLT CP at 6 months and 83.9% and 80.8% at 12 months (p= 0.31 and 0.68, respectively). Morphologic changes were not correlated with LDL-C variations over time.

### Plaque regression

Mean baseline stenosis was 57±5% in both groups (Table III). At 6 months, PR occurred in 30.4% and 20.5% in the ELLT and LLT group, respectively (p=0.17). At 12 months, among 162 CP (that is, 160 with 12 month-DUS and the 2 plaques who underwent CR, then with no PR), PR occurred in 63 plaques (39%), without statistical difference between the two groups: 43% in the ELLT and 35.1% in the LLT group (p=0.42). No statistical differences between the two groups were found regarding PW and PQ (Figure 2). No correlation between quantitative changes and LDL-C variations was observed over time.

### Primary endpoint

The primary endpoint (MS at 6 months and/or PR at 12 months) was met in 65 CP overall: 39 out of 88 ELLT plaques (44.3%) versus 26 out of 74 (35.1%) LLT plaques (p=0.26)

### Patients

#### Lipid-lowering therapy

At baseline, statin therapy was ongoing in 79% patients, but only 29% were on full-dose statin and 12.1% on full-dose statin+ezetimibe. All were upgraded to 20 mg rosuvastatin + 10 mg ezetimibe. Within 12 months, 8 patients stopped statin (4 in each group), 15 patients (7 ELLT and 8 LLT) reduced statin dose, and 4 stopped ezetimibe (2 in the ELLT and 2 in the LLT group).

Reasons for statin discontinuation were statin-associated muscle symptoms (SAMS) in all cases except: 1 LLT patient, who developed myasthenia gravis, 1 ELLT patient in whom creatinine increased, and 1 ELLT patient a whose statin had been stopped by the primary physician, who judged LDL-C values (39 mg/dL) “too low”.

Reasons for statin dose reduction were SAMS in all cases, except 2 ELLT patients: creatinine elevation occurred in one, “too low” LDL-C values (8 mg/dL) in the other. Both patients who stopped/ reduced statin due to creatinine increase had an LDL-C <20 mg/dL. No CK elevation>4 ULN was observed.

After 6 months, 6 patients stopped evolocumab, either for missed prescription or unwillingness to continue it. No allergic reactions to evolocumab were reported.

#### LDL-C

Baseline LDL-C was 126±30 mg/dL, with no difference between the two groups; total cholesterol was slightly higher in the ELLT group (Table IV). Overall, LDL-C was more than halved at 6 months. At 6 and 12 months, LDL-C was significantly lower in the ELLT group (32±35 versus 64±30, p=0.01 and 34±34 versus 64±30, p=0.001, respectively). At 12-month, the LDL-C target≤55 mg was achieved in 83.4% with ELLT *versus* 43.1% with LLT (p=0.0001). Triglycerides were significantly lower in the ELLT group after 6 months; no significant HDL-C variations occurred between the two groups. (Table IV).

**Table IV.** Lipid profile variations.

| Outcome | Timepoint | N | Overall |  | N | ELLT |  | N | LLT |  |  | Between Groups | Within-Groups | Interaction |
| --- | --- | --- | --- | --- | --- | --- | --- | --- | --- | --- | --- | --- | --- | --- |
|  |  |  | Values | Change |  | Values | Change |  | Values | Change | Mean (95% CI) | p |  |  |
| LDL-C | Baseline | 165 | 125.95 ± 29.61 |  | 83 | 127.54 ± 26.94 |  | 82 | 123,24 ± 30.67 |  |  |  |  |  |
|  | 6 months | 150 | 47.41 ± 38.58 | -76.48 ± 40.92 | 78 | 32.08 ± 35.12 | -95.98 ± 38.44 | 72 | 64.25 ± 35.22 | -55.05 ± 32.02 | 40.94 (29.42,52.45) | 0.01 | 0.001 | 0.001 |
|  | 12 months vs 6 months | 150 | 48.12 ± 35.52 | 0.39 ± 31.45 | 78 | 33.79 ± 34.29 | 1.71 ± 29.62 | 72 | 63.65 ± 30.04 | -1.09 ± 33.50 | -2.80 (-13.02,7.42) | 0.001 | 0.9043 | 0.5894 |
|  | 12 months vs baseline | 150 | 48.12 ± 35.52 | -75.60 ± 41.36 | 78 | 33.79 ± 34.29 | -94.27 ± 37.77 | 72 | 63.65 ± 30.04 | -55.38 ± 35.29 | 38.89 (27.07,50.71) | 0.001 | 0.001 | 0.001 |
| LDL-C <70 mg/dL | Baseline | 165 | 0 |  | 83 | 0 |  | 82 | 0 |  |  |  |  |  |
|  | 6 months | 150 | 118 (79) | 118 | 78 | 67 (86) | 67 | 72 | 51 (71) | 51 |  | 0.029 | 0.001 | 0.001 |
|  | 12 months vs 6 months | 150 | 118 (79) | 0 | 78 | 68 (87) | 1 | 72 | 50 (69) | -1 |  | 0.001 | 0.457 | 0.473 |
|  | 12 months vs baseline | 150 | 118 (79) | 118 | 78 | 68 (87) | 68 | 72 | 50 (69) | 50 |  | 0.01 | 0.001 | 0.001 |
| LDL-C <55 mg/dL | Baseline | 165 | 0 |  | 83 | 0 |  | 82 | 0 |  |  |  |  |  |
|  | 6 months | 150 | 96 (64) | 96 | 78 | 63 (81) | 63 | 72 | 33 (46) | 33 |  | 0.01 | 0.0001 | 0.022 |
|  | 12 months vs 6 months | 150 | 92 (61) | -4 | 78 | 65 (83) | 2 | 72 | 31 (43) | -2 |  | 0.022 | 0.761 | 0.702 |
|  | 12 months vs baseline | 150 | 92 (61) | 92 | 78 | 65 (83) | 65 | 72 | 31 (43) | 31 |  | 0.0001 | 0.001 | 0.005 |
| Total Cholesterol | Baseline | 164 | 198.01 ± 39.42 |  | 83 | 203.76 ± 33.19 |  | 81 | 192.11 ± 44.35 |  |  |  |  |  |
|  | 6 months | 149 | 118.43 ± 45.47 | -79.34 ± 48.82 | 78 | 100.82 ± 41.15 | -103.641 ± 42.47 | 71 | 137.78 ± 42.23 | -52.63 ± 40.90 | 51.01 (37.48,64.53) | 0.0140 | 0.001 | 0.001 |
|  | 12 months vs 6 months | 150 | 116.6 ± 40.53 | -2.15 ± 34.18 | 78 | 102.10 ± 41.42 | 1.28 ± 34.65 | 72 | 132.31 ± 33.26 | -5.92 ± 33.49 | -7.20 (-18.25,3.86) | 0.00001 | 0.4088 | 0.2002 |
|  | 12 months vs baseline | 150 | 116.6 ± 40.53 | -80.91 ± 48.12 | 78 | 102.10 ± 41.42 | -102.359 ± 42.12 | 72 | 132.31 ± 33.26 | -57.67 ± 43.38 | 44.69 (30.89,58.49) | 0.0398 | 0.001 | 0.001 |
| HDL-C | Baseline | 164 | 49.80 ± 13.85 |  | 83 | 50.24 ± 13 |  | 81 | 49.36 ± 14.75 |  |  |  |  |  |
|  | 6 months | 149 | 51.79 ± 13.49 | 1.93 ± 10.95 | 78 | 51.80 ± 13.11 | 1.13 ± 7.62 | 71 | 51.79 ± 13.99 | 2.82 ± 13.71 | 1.69 (-1.86,5.24) | 0.9848 | 0.0296 | 0.3487 |
|  | 12 months vs 6 months | 150 | 51.08 ± 12.28 | -0.73 ± 6.80 | 78 | 51.39 ± 11.92 | -0.41 ± 7.08 | 72 | 50.75 ± 12.74 | -1.09 ± 6.50 | -0.67 (-2.88,1.53) | 0.8845 | 0.1829 | 0.5470 |
|  | 12 months vs baseline | 150 | 51.08 ± 12.28 | 1.29 ± 11.07 | 78 | 51.39 ± 11.92 | 0.72 ± 6.93 | 72 | 50.75 ± 12.74 | 1.90 ± 14.30 | 1.19 (-2.40,4.77 | 0.8798 | 0.1504 | 0.5144 |
| Tryglicerides | Baseline | 166 | 126.57 ± 52.33 |  | 83 | 130.98 ± 48.34 |  | 82 | 122.18 ± 56.33 |  |  |  |  |  |
|  | 6 months | 149 | 99.12 ± 57.52 | -24.94 ± 55.10 | 78 | 89.59 ± 58.98 | -40.32 ± 58.36 | 71 | 109.59 ± 54.38 | -8.04 ± 46.03 | 32.28 (15.15,49.41) | 0.3416 | 0.001 | 0.03 |
|  | 12 months vs 6 months | 148 | 94.82 ± 44.93 | -4.13 ± 47.46 | 78 | 86.60 ± 43.34 | -2.99 ± 53.87 | 70 | 103.97 ± 45.20 | -5.4 ± 39.46 | -2.41 (-17.90,13.08) | 0.0122 | 0.2863 | 0.7586 |
|  | 12 months vs baseline | 148 | 94.82 ± 44.93 | -28.88 ± 54.35 | 78 | 86.60 ± 43.34 | -43.31 ± 50.08 | 70 | 103.97 ± 45.20 | -12.8 ± 54.75 | 30.51 (13.48,47.54) | 0.3343 | 0.001 | 0.05 |

Data are presented as numbers (percentage), or mean ± SD, as appropriate
Numeric changes are the net algebraic changes and were calculated considering only available data
Between-groups: effect of time periods (6months versus baseline, 12 months versus 6 months, 12 months versus baseline) across the two groups
Within-group: effect of therapy across the time periods in each group; i.e., effect of EOLLT across the time periods in the EOLLT group
Interaction: indicates whether the effect of therapy depends on the time periods
ELLT= evolocumab on top of OLLT; LLT= optimal lipid-lowering therapy

#### Secondary endpoint

The 12-month LDL-C absolute and % change as compared to baseline were -94.27 mg/dL and - 73.5% with ELLT and -55.38 mg/dL and -48.2% with LLT (p=0.0001 for both).

#### Adverse events

Overall, adverse events occurred in 16 patients at 1 year (Table V); one patient developed myasthenia gravis, 2 patients died (one due to cancer: glioblastoma). Additionally, 2 ischemic strokes occurred, 3 myocardial infarctions, 2 CR, 3 PCI, and 5 peripheral revascularizations. Three patients experienced more than one event. In particular, MAVE (Figure 3) occurred in 2 patients treated with ELLT (2.4%) and 12 (14.6%) treated with LLT (p= 0.005). None of these patients with MAVE declared modification in the assigned therapy before the event. In addition, one of the patients who withdrew trial consent after randomization underwent sCR within 6 months; he was taking rosuvastatin 10 mg. No correlation of MAVE with LDL-C values was seen. At multivariate analysis, which included other than carotid PAD, CAD, and the study treatment, the absence of Evolocumab was the only independent predictor of MAVE (OR 6.96, 95% CI 1.5-32.5, p = 0.014; Hosmer-Lemeshow p=0.73). As a sensitivity analysis, given the limited number of events (n=14), Firth penalized logistic regression confirmed that the absence of evolocumab was the only independent predictor of MAVE (OR 5.68, 95% CI 1.62–30.3, p=0.005).

**Table V.** Adverse events and characteristics at enrollment.

| # patient | Event | Months | Treatment | Gender | Smoking | Diabetes | CAD | PAD | LDL-C (mg/dL) | Antiplatelet | Full-dose statin | Carotid Morphological Type | Carotid stenosis (%) | Carotid PSV (cm/s) |
| --- | --- | --- | --- | --- | --- | --- | --- | --- | --- | --- | --- | --- | --- | --- |
| 1 | MI and stroke | 3 | LLT | F | 0 | 0 | 0 | 0 | 198 | 0 | 0 | IV | 65 | 105 |
| 2 | pLR; PCI | 6; 11 | LLT | M | 1 | 1 | 1 | 1 | 114 | 1 | 1 | III | 60 | 140 |
| 3 | pLR | 4 | LLT | F | 1 | 0 | 0 | 1 | 197 | 1 | 0 | IV | 50 | 220 |
| 4 | Stroke | 3 | LLT | M | 1 | 0 | 1 | 0 | 103 | 1 | 1 | III | 50 | 70 |
| 5 | pCR | 2 | LLT | M | 1 | 0 | 0 | 1 | 168 | 1 | 0 | III | 50 | 130 |
| 6 | EVAR + sLR | 6 | LLT | M | 1 | 0 | 0 | 1 | 111 | 1 | 0 | IV | 50 | 60 |
| 7 | MI | 9 | LLT | M | 0 | 1 | 0 | 0 | 109 | 1 | 0 | II | 55 | 70 |
| 8 | PCI | 5 | ELLT | M | 0 | 1 | 0 | 0 | 128 | 1 | 1 | II | 60 | 120 |
| 9 | pLR | 10 | LLT | F | 0 | 1 | 0 | 1 | 111 | 1 | 0 | I | 65 | 220 |
| 10 | MI | 5 | LLT | M | 0 | 0 | 0 | 0 | 157 | 1 | 0 | II | 65 | 150 |
| 11 | sCR | 11 | ELLT | M | 1 | 0 | 0 | 0 | 121 | 1 | 0 | I | 70 | 280 |
| 12 | PCI | 9 | LLT | F | 1 | 1 | 1 | 0 | 104 | 1 | 1 | III | 60 | 105 |
| 13 | sLR | 10 | LLT | F | 0 | 1 | 0 | 0 | 117 | 1 | 0 | III | 55 | 130 |
| 14 | Cardiac death | 10 | LLT | M | 0 | 1 | 0 | 0 | 179 | 1 | 0 | I | 50 | 100 |
| 15 | Non-cardiac death | 11 | LLT | M | 1 | 0 | 1 | 1 | 100 | 1 | 1 | I | 55 | 130 |
| 16 | Myasthenia gravis | 7 | LLT | M | 0 | 1 | 0 | 0 | 108 | 0 | 1 | III | 55 | 105 |

Data are presented as numbers (percentage), or mean ± SD, as appropriate
ELLT= evolocumab on top of LLT. LLT= lipid-lowering therapy
Prefixes: p =percutaneous, s=surgical
CR= carotid revascularization
EVAR= Endovascular Aortic Repair
LR= limb revascularization
MI = Myocardial Infarction
PCI= percutaneous Coronary Intervention
PSV = Peak systolic velocity

**Figure 3.**
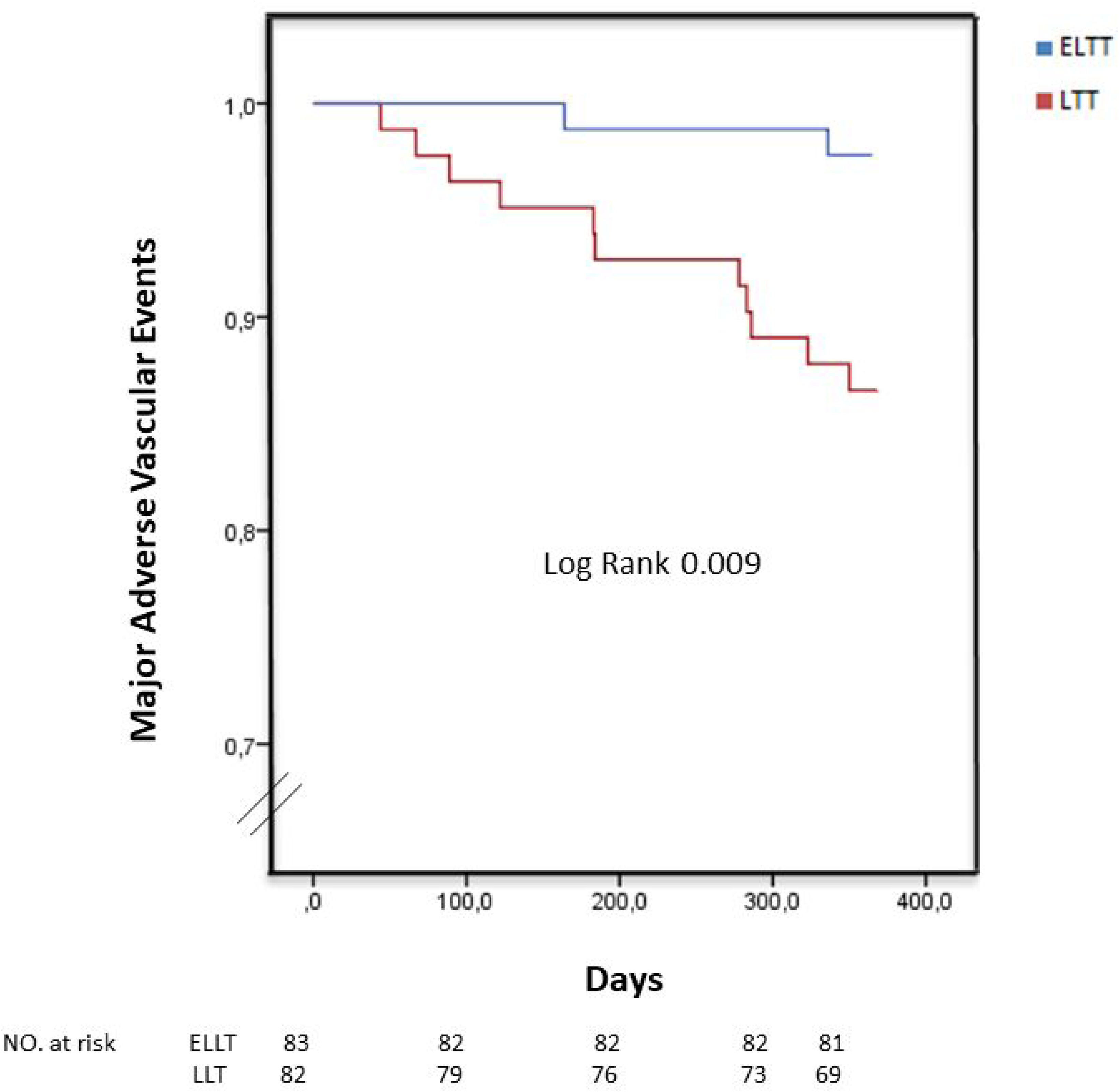
Major adverse vascular events (cardiac death, stroke, myocardial infarction, carotid or coronary or peripheral revascularization) at one year. LLT= optimal lipid-lowering therapy; ELLT= evolocumab on top of LLT.

## DISCUSSION

The CARUSO trial is the largest study comparing ELLT and LLT in patients with CP≥50% and LDL-C>100 mg/dL. Besides confirming the persistent undertreatment of patients with carotid artery disease in the current era, our data,

1. Highlight the dynamic nature of CP morphology, and shows that a low-risk morphological type at baseline does not ensure subsequent stability. Notably, the low-risk type III subgroup proved to be the most dynamic, underscoring the importance of strict LDL-C control even when baseline morphology appears benign;
2. Demonstrate that both ELLT and optimized LLT are associated with MS and PR, with most changes occurring within the first six months. The primary composite endpoint of 6-month MS and 12-month PR was numerically higher with ELLT, without reaching statistical significance. However, evolocumab significantly prevented transitions from low-risk to high-risk morphological categories compared with LLT;
3. Indicate that the 1-year rate of MAVE was reduced sevenfold with evolocumab, which emerged as the only independent predictor.

### 1. Morphologic and quantitative changes

#### Entire cohort

Overall, 6-month-MS and 12-month-PR occurred with both ELLT and LLT, as in previous studies.^13^ Most morphologic and quantitative changes occurred in the first 6 months, and 12-month-to-baseline differences were mostly driven by changes at 6 months. This can have two, non-mutually exclusive, explanations.

a. In both the LLT and ELLT groups, a drug-action plateau is achieved within 6 months, after which a longer time is needed to achieve further passivation. In the ELLT group, high-risk plaque numerically decreased at 6 months, then increased at 12 months, and less stenosis reduction>5% occurred between 6 and 12 months compared to the period from baseline to 6 months. This possible plateau, together with the fact that low-risk CP may progress to high-risk types, and that carotid atherosclerosis continues to progress in >20% of patients, despite achieving LDL-C targets ^17^ represents a call to prompt therapeutic action independently of the CP morphology and stenosis.
b. Adherence to therapy reduces after 6 months, and, likewise, favorable CP changes reduce. Missed compliance with LLT is a common event in clinical practice. However, it might also occur with evolocumab, since its availability in Italy is limited by a special therapeutic prescription that is restricted to non-GP specialists and renewed every 6 months. This, together with the low awareness regarding primary prevention in patients with carotid disease, and the persistent false belief that very low LDL value may be dangerous despite available evidence,^1, 18, 19^ may further discourage adherence to ELLT.

#### ELLT versus LLT

Evolocumab was associated with more favorable morphological and quantitative changes than LLT: 6-and 12-month MS was more frequent (10% *versus* 0; 8% *versus* 5%), and MD was more than halved (4% *versus* 10%; 6% *versus* 15%). In particular, while all plaques that shifted from high to low-risk achieving 6-month-MS were ELLT CP, those who shifted from low to high-risk types at 6 and 12 months were mostly LLT. These shifts also indicate that a baseline low-risk type does not warrant MQ.^20, 21^

It is important to note that the different baseline number of high-risk CP in the 2 groups did not influence the morphological component of the primary endpoint. Indeed, morphological stabilization is defined as the shift from high-risk to low-risk plaque type. Only high-risk plaques can shift to low risk, and the proportion of high-risk plaques that shifted to low-risk types achieved morphological stabilization. Therefore, even if the baseline number of high-risk plaques was different between groups, what matters is the proportion of shifts.

Six- and 12-month PR were numerically higher in the ELLT group too.

The association of evolocumab with plaque passivation and regression is intriguing. So far, evidence has been limited to case reports, retrospective studies and very small randomized studies. A retrospective DUS study showed that evolocumab in statin-treated patients significantly reduced the 12-month increase in carotid intima-media thickness, as compared to the previous 12 months with statins only.^22^ Randomized studies with magnetic resonance imaging (MRI) showed either higher 6-month-PR^14^ or 12-month-MS^13^ with ELLT compared with statins, or no statistical difference.^23^

There are several possible reasons for the missed statistical significance regarding the primary endpoint in the CARUSO study.

### I. Therapeutic delta

This is the only available study on CP in which LLT was uniformly uptitrated at enrollment. At baseline, only 29% of patients were on full-dose statin, yet another confirmation of the widespread undertreatment of patients with PAD,^24, 25^ despite guidelines recommendation.^5^ In all patients, the baseline lipid-lowering therapy was uptitrated to an optimized LLT (rosuvastatin 20 mg + ezetimibe 10 mg) to have the same background therapy for evolocumab and a uniform control arm, rather than a variety of different baseline therapies (including no treatment in 21%, Table II).

Rosuvastatin 20 mg is considered a high-intensity lipid-lowering treatment, and the expected average reduction in LDL-C levels in combination with ezetimibe in naïve patients is 60%,^5^. therefore it is correct to define it as optimal lipid-lowering therapy. Moreover, the combination of rosuvastatin 20 mg and ezetimibe offers a good profile of tolerability with respect to side effects. This uptitration almost halved 6-month-LDL-C in the LLT arm, but possibly reduced the expected therapeutic delta between ELLT and LLT and/or prolonged the time needed to see differences between the two groups. Leaving patients on their “naive” LLT would have likely increased the relative rate of ELLT-induced MS and PR, possibly reaching statistical significance of the primary endpoint, but would not have been ethical. This is even truer with hindsight, after having observed the higher 1-year MAVE rate in the LLT group, which occurred despite the therapeutic upgrading.

The same concept applies to stenosis degree, whereas the impact of ELLT on PR could have been higher if basal stenoses were more critical. Indeed, the only study^14^ that showed significant 6-month-PR (defined as lipid-rich necrotic core (LRNC) changes) with evolocumab as compared to statins included stenosis well higher than CARUSO’s. Having >70% as threshold for considering carotid endarterectomy,^26^ the inclusion of higher-degree stenosis would have encountered the reluctance of vascular surgeons to wait for PR, although it has been observed.^12^ Notwithstanding, in our 50-70% stenosis range, 12-month-PR was observed in 43% CP with evolocumab.

### II. DUS

Plaque evolution was monitored with DUS, which is the most available, user-friendly, and cost-effective imaging method, and remains the first-line diagnostic exam in clinical practice. However, DUS has a reduced sensitivity as compared with MRI, especially regarding CP composition and early detection of variations. A retrospective study evaluated carotid total plaque area (TPA) with DUS in 131 patients during a period of 6±4 years and showed a decrease in TPA, particularly during the first 3 years with PCSK9-inhibitors.^27^ However, TPA was defined as the sum of the areas of all plaques between the clavicle and the angle of the jaw in the right subclavian and common, internal, and external carotid arteries, while only internal CP were considered in our study.

Previous studies with MRI showed favorable CP modifications with statins.^28–30^ More recent MRI studies evaluated the impact of evolocumab on top of LLT by examining changes in CP LRNC.^23^ ^14^ One study included 33 CARUSO-like patients with stenosis≥50% and LDL-C ≥1.8 mmol/L (70 mg/dL). ^23^At 12 months, LRNC was numerically but not statistically reduced with evolocumab. In this study, no differences in stenosis and plaque type were detected at DUS.

Another study^14^ including 63 patients, showed that LRNC volume in the evolocumab arm (mean baseline stenosis 76.6%) was significantly reduced as compared to standard LLT (mean baseline stenosis 81.1%) at 6 months, with an NNT to prevent progression of 3.

Other imaging techniques may also be useful: in a randomised, placebo-controlled trial alirocumab reduced CP inflammation detected using PET/CT in statin-intolerant patients.^31^

However, as MRI they are not always available. Contrast-enhanced US may be a good compromise. It was used in 41 patients with premature CAD to explore the 1-year effect on CP evolution of evolocumab on top of statins as compared with placebo.^13^ Both evolocumab and statins reduced carotid intraplaque neovascularization (IPN) and max plaque height (MPH), with no difference in 1-year MPH reduction (that means PR) between the two groups, but greater IPN reduction (that means MS) with evolocumab.

### III. Sample size and Timeline

All studies on CP following therapy with PCSK9-inhibitors have been limited to small numbers. Although CARUSO is the largest study so far, the relative small sample size may be an additional reason for the lack of statistical significance regarding the primary endpoint. Another important reason is that one year may not be enough to observe significant PR,^13^ as also shown with statins.^28^ While ELLT may block natural history towards MD, the achievement of MS possibly requires a more than 6 months, as shown by a single-arm MRI study including 27 patients where reduction of both CP lipid core and neovasculature by alirocumab were seen at 12 months.^32^ Furthermore, starting from a mean stenosis of 57%, an even longer time may be required to observe PR.

### 2. LDL-C

In line with literature data,^7, 8^ 12-month LDL-C was reduced by 74% in the ELLT arm, as compared with 48% in the LLT arm (p=0.0001). There was no correlation between LDL-C and CP modification, as previously observed.^28, 30, 32, 33^ Also, a metanalysis showed that PCSK9-inhibitors promoted stroke reduction independently of LDL-C decrease.^34^ Therefore, other factors may be involved in CP evolution, like platelet-derived growth factor.^35^ Furthermore, a direct effect of evolocumab on atherosclerosis pathophysiology is implicated, thus explaining CP passivation. Indeed, PCSK9-inhibitors reduce by 20–30% lipoprotein(a)^8, 36^ and block PCSK9, which are both directly involved in atherosclerotic plaque formation and growth regardless of LDL-C.^37,38, 39^ They also reduce inflammation, shear stress/reactive oxygen species, triglycerides, and platelet aggregation.^9, 40^ Moreover, LDL-C change could not reflect MS and/or PR in the single patient since CP evolution is a multifactorial event, influenced by smoking status, hypertension control, antiplatelet therapy, diabetes, and other atherogenic factors. Therefore, in a patient with a heap of cardiovascular risk factors, LDL-C goals needed to achieve MS/PR could be even lower than those currently recommended.

### 3. Cerebro and Cardiovascular adverse events

Carotid stenosis is a known independent predictor of MAVE,^41^ despite best medical therapy, including LLT and anti-platelets.^42^ Previous studies showed that PCSK9-inhibitors reduce stroke and TIA in patients with CAD.^8, 43^

In our study, evolocumab was associated with a sevenfold risk reduction of 1-year atherosclerotic events, with an NNT to prevent one MAVE of 8. The impact of ELLT on atherosclerosis passivation was translated into the clinical scenario, suggesting a systemic effect beyond that on the CP. Otherwise, CP macroscopic characterization/ quantification with DUS may not provide complete information about the systemic antiatherosclerotic effects of evolocumab. The early and striking clinical impact of ELLT underlines both the drug efficacy and the ultra-high atherosclerotic risk of the study cohort. Thus, the presence of carotid stenosis≥50% with LDL-C not at target identifies patients who would significantly benefit from evolocumab regardless of the morphological type and stenosis degree.^44^ In this view, ELLT should be administered also in patients who are already candidates for invasive carotid treatment. Morphologic and quantitative worsening should not discourage ELLT use or accelerate intervention decisions, while watchful waiting, in the absence of symptoms, may be wiser.^12, 45^ Furthermore, those with high-risk CP have a higher propensity to periprocedural embolization and stroke after invasive treatment, which may be reduced, as shown with statins.^46^ Dedicated studies are needed to explore these concepts. However, the results of the recent VESALIUS-CV trial, although only 10% of patients had cerebrovascular disease, confirm the efficacy of evolocumab in primary prevention in high-risk populations.^47^

#### Limitations

The main study limitation is the relatively small sample size, although it is the largest to date. Future larger studies including high-risk CP only or higher-degree stenosis may achieve further evidence. The absence of an independent core lab and clinical event committee may be another limitation. However, the data manager and statistician were blinded to the assigned treatment, and the agreement of the two vascular surgeons in labelling CP morphology and stenosis was very high (k coefficient =0.9). Lastly, the analyses followed an intention-to-treat approach, and we might not have recorded all the changes in the background therapy during the study period.

#### Perspectives

Future larger studies are warranted to validate our findings, assess long-term adherence to therapy, and identify subgroups with a higher probability of achieving MS and PR. This would be important to promote therapy cost-effectiveness. Considering the striking prognostic impact, ELLT should be considered an integral part of the management of patients with CP, regardless of the indication for carotid intervention.

## CONCLUSIONS

The CARUSO trial is the largest randomized study testing the effect of evolocumab on carotid plaque morphological stabilization and regression in patients with carotid stenosis≥50% and LDL-C≥100 mg/dL. Compared to LLT, ELLT was associated with numerically, but not statistically, higher 6-month MS and/or 12-month PR. In the LLT group, changes from low to high-risk types, LDL-C, and MAVE were significantly higher. The absence of evolocumab was the only independent predictor of MAVE. According to our results, ELLT might become the standard treatment for patients with ≥50% stenosis and LDL-C>100 mg/dL.

### Sources of funding

No funding to declare

### Disclosures

No conflict of interest to declare

### Data availability statement

The data supporting the study findings are available from the corresponding author upon reasonable request.

## FIGURE LEGEND

Central Illustration. Methods and Results of the CARUSO study.

CP = carotid plaque

